# Digital Engagement of Neurocutaneous Syndrome Communities on Facebook: A Descriptive Study

**DOI:** 10.1101/2024.09.18.24313923

**Authors:** Donia Momen, Kushagra Patel, Logan Muzyka, Michael DeCuypere, Sandi K. Lam, Jeffrey S. Raskin

**Author notes:** **Corresponding Author:** Jeffrey S. Raskin, MS MD. Funding: There were no funding sources to disclose. Disclosures: Nothing to disclose.

## Abstract

**Introduction:** Social media has become a platform for healthcare providers and parents to communicate about common and rare pediatric disorders. Neurocutaneous syndromes including Tuberous Sclerosis Complex, Neurofibromatosis, and Sturge-Weber syndrome require lifelong monitoring and complex treatments. Clinician directed information is supplemented by online support groups, non-profit fundraising organizations, and educational programs through various Facebook features.

**Objective:** The objective of this study is to explore the socialization patterns of individuals affected by neurocutaneous syndromes as they interact and engage within various Facebook forums.

**Methods:** Public accounts on Facebook were analyzed throughout August 2022. The search terms used were “Sturge-Weber Syndrome”, “Sturge Weber”, “SWS”, “Tuberous Sclerosis”, “Tuberous Sclerosis Complex (TSC)”, and “Neurofibromatosis”, “Neurofibromatosis 1 (NF1)”, and “Neurofibromatosis 2 (NF2)”. Facebook accounts were analyzed on the basis of page ownership and video content.

**Results:** Searching Sturge-Weber syndrome (SWS) yielded 99 Facebook pages and 37 Facebook videos. Tuberous sclerosis resulted in 100 Facebook pages and 139 Facebook videos. Neurofibromatosis yielded 102 Facebook pages and 129 Facebook Videos. Patient released stories comprised 51% of the Facebook videos (157/305), educational videos comprised 37% (112/305), and 12% (36/305) related to the syndromes’ awareness day. Facebook Pages were managed by nonprofit organizations 37% (111/301), community support groups 36% (108/301), and patients creating a “private blog” 23% (70/301), and physicians 4% (12/301). Figure 1 shows a breakdown of the Facebook page ownership and video content of neurocutaneous syndromes.

**Conclusion:** This study describes the online discussion of neurocutaneous syndromes through social media. The benefits of anonymized discussion for patients and families are many. Physicians manage only 4% of the Facebook pages and this may represent an opportunity to further disseminate medical advice in the future.

## Introduction

In today’s digital age, social media platforms have evolved into powerful tools for healthcare providers and parents to disseminate information and communicate about disorders. Since COVID-19 lockdown, the social media platforms have become the mainstream channels for acquiring information and finding social support.^1^ This has been observed through personal searches, in individuals affected by neurocutaneous syndromes, which encompass conditions such as tuberous sclerosis complex, neurofibromatosis, and Sturge-Weber syndrome. These syndromes necessitate lifelong monitoring and nuanced treatment plans. Healthcare professionals deliver clinician-directed information which is often supplemented with online social media resources. Online support groups, non-profit fundraising organizations, and educational programs facilitated through platforms like Facebook, provide a collaborative and multifaceted approach to support individuals and families facing neurocutaneous syndromes.^2^

Facebook is a popular online platform that fosters communities of like-minded individuals who can share their medical experiences, share their stories, seek advice, and find emotional support by forming ‘groups’ and making ‘pages’.^6^ This platform also enables users to raise awareness, advocate for medical causes, and share credible information.^6^

Descriptive analysis of FB use patterns among patients and physicians has not yet been conducted. This study aims to assess how individuals affected by neurocutaneous syndromes socialize through various Facebook Pages and related videos, as well as to understand how these platforms are used to support and empower those impacted by these conditions.

### Methods

We conducted an analysis of public Facebook accounts from January 2022 to December 2023 to investigate the presence and content related to specific neurocutaneous syndromes. The search terms used were “Sturge-Weber Syndrome”, “Sturge Weber”, “SWS”, “Tuberous Sclerosis”, “Tuberous Sclerosis Complex (TSC)”, and “Neurofibromatosis”, “Neurofibromatosis 1 (NF1)”, and “Neurofibromatosis 2 (NF2)”. Facebook accounts were analyzed by comparing content from Pages dedicated to the syndromes as well as videos posted related to these conditions.

Facebook Pages and Videos were further broken down based on the content they provided. Pages included four categories: “non profit organizations”, “community support group”, “patient private blog”, or “physician’s page”. There were three video categories: “patient related stories”, “educational videos”, and “awareness day”. We determined the number of Videos / Pages and the total within each category.

Subsequently, the top Page for each search term was further analyzed based on public pages and the number of followers, with a minimum threshold of one-thousand followers. The “SWS Awareness Day” Page (9,500 followers), “Neurofibromatosis Network” Page (17,000 followers, and “Tuberous Sclerosis Association” Page (4,600 followers qualified as the top Pages. The “Neurofibromatosis Network” Page was composed mainly of NF1 context; however, the Page also included NF2, making it a sum of both NF1 and NF2.

Sub categories were used to determine the context of the content posted on each Page: “asking for social / health advice”, “health education / awareness”, “personal stories”, “page engagement posts”, “support groups”, “other”.^12^ “Other” accounted for anything that did not fit into the other categories, including research study recruitment, participation or funding and a reality medicine tv show advertisement.

We maintained ethical considerations by focusing solely on public accounts while respecting user privacy and confidentiality. Each post was evaluated by the primary researcher and placed into the most suitable and accurate category. A second researcher independently confirmed placement. Interrater reliability was 100%, demonstrating full agreement among both researchers. All data collected and analyzed adhered to Facebook’s terms of service.

IRB approval was not obtained due to the use of publicly available data.

## Results

A total of 422 Facebook Videos were identified related to either Sturge-Weber Syndrome (n=91, 22%), Neurofibromatosis (n=136, 32%) or Tuberous Sclerosis (n=195, 46%). Table 1 reflects that each grouped syndrome had higher *Patient Stories* videos (SWS: 48%, NF: 47%, TS: 50%), then *Educational Videos* (SWS: 43%, NF: 40%, TS: 41%), and lastly, *Awareness Day* (SWS: 9.0%, NF: 13%, TS: 9.0%).

**Table 1.** Overview of Facebook Videos with number of posts subcategorized comparing 3 neurocutaneous syndromes.

| Facebook Videos |  |  |  |  |  |  |
| --- | --- | --- | --- | --- | --- | --- |
| Categories |  | Sturge-Weber Syndrome |  | Neurofibromatosis |  | Tuberous Sclerosis |
| Patient Stories |  | 44 (48%) |  | 64 (47%) |  | 98 (50%) |
| Educational Videos |  | 39 (43%) |  | 54 (40%) |  | 80 (41%) |
| Awareness Day |  | 8 (9.0%) |  | 18 (13%) |  | 17 (9.0%) |
| Total (422) |  | 91 (22%) |  | 136 (32%) |  | 195 (46%) |

Facebook Pages were very consistent across the studied syndromes (SWS: 30%, NF: 33%, TS: 37%), with a total of 344 Pages identified. Table 2 presents the frequency of categorized pages, with *Non-Profit Organization* (SWS: 36%, NF: 42%, TS: 33%) the most represented, followed by *Support Groups* (SWS: 33%, NF: 40%, TS: 31%), *Private Blogs* (SWS: 20%, NF: 13%, TS: 29%) and *Physicians Pages* (SWS: 11%, NF: 5.0%, TS: 7.0%).

**Table 2.** Overview of Facebook Pages subcategorized posts based on different content comparing all 3 neurocutaneous syndromes.

| Facebook Pages |  |  |  |  |  |  |
| --- | --- | --- | --- | --- | --- | --- |
| Categories |  | Sturge-Weber Syndrome |  | Neurofibromatosis |  | Tuberous Sclerosis |
| Non-Profit Organizations |  | 37 (36%) |  | 48 (42%) |  | 42 (33%) |
| Support Groups |  | 34 (33%) |  | 46 (40%) |  | 39 (31%) |
| Private Blogs |  | 21 (20%) |  | 15 (13%) |  | 36 (29%) |
| Physicians Pages |  | 11 (11%) |  | 6 (5.0%) |  | 9 (7.0%) |
| Total (344) |  | 103 (30%) |  | 115 (33%) |  | 126 (37%) |

Top Page analysis for each neurocutaneous syndrome was performed from January 2022 to December 2023 and reflected different subcategorization makeup (Table 3): SWS Awareness Day Page emphasized *Personal Stories* (n=168, 74.0%), NF Network and TS Association emphasized*Page Engagement Posts* (n=135, 44.1% ; n=198, 54.1%, respectively. TS Association was the most active Page accounting for 366 posts (40.7%) out of the total of 899.

**Table 3.**
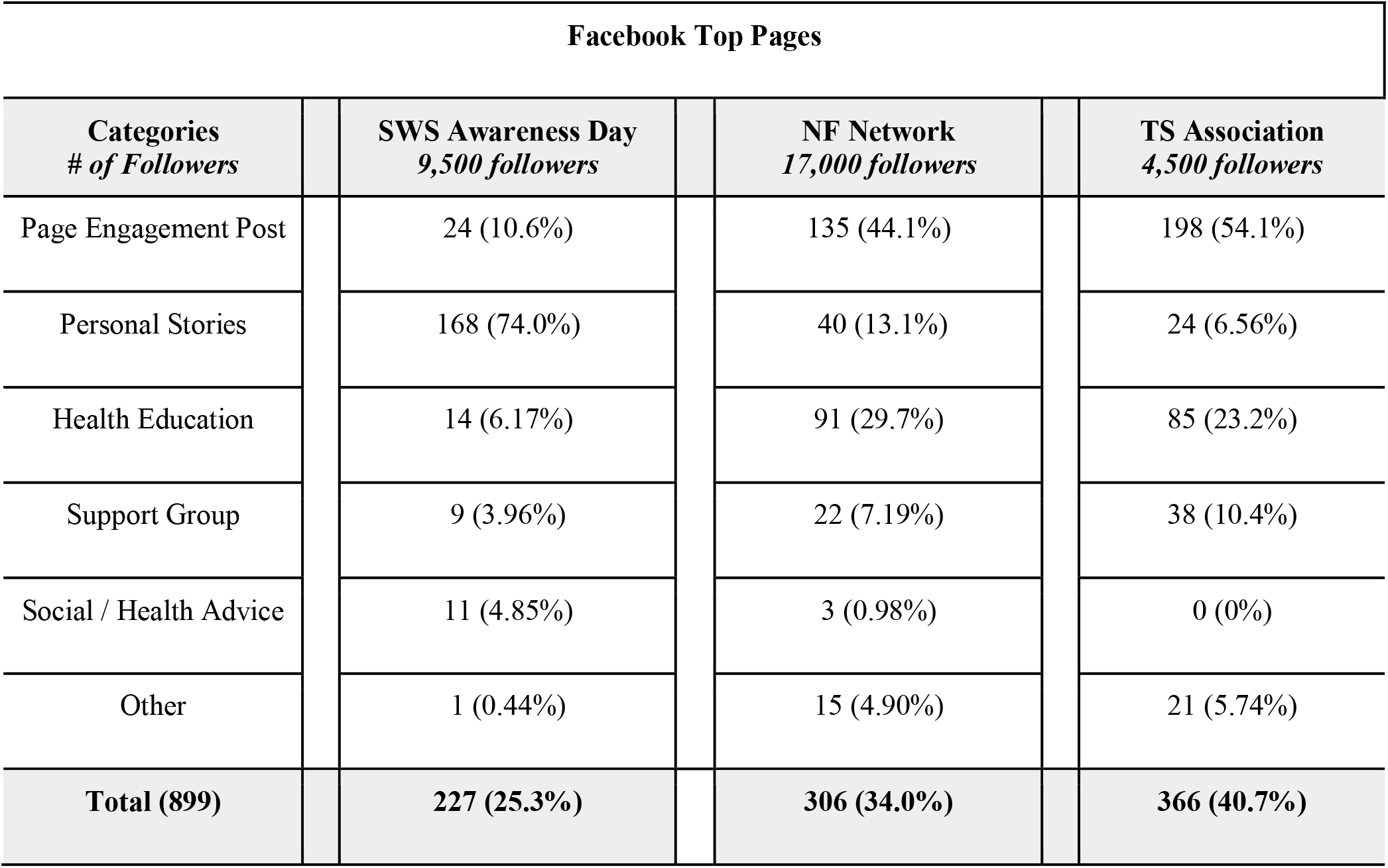
Subcategorization of the top Facebook Page for each neurocutaneous syndrome based on the following count. Percentages are rounded to 3 significant figures and may not add exactly to 100%.

## Discussion

We aimed to perform a descriptive analysis of the dimensions within social media use by three groups of neurocutaneous syndromes, Sturge Weber, Neurofibromatosis, and Tuberous Sclerosis.

Primarily, our study revealed that a significant portion of video content — 91% of SWS, 87% of NF, and 91% of TSC — was defined by compelling patient narratives, offering personal insights and experiences, and educational videos, dedicated to disseminating awareness concerning disease symptoms and treatment outcomes. This dominance of patient stories and educational content underscores their role in shaping the discourse surrounding neurocutaneous disorders on Facebook. The remaining 10% of videos predominantly highlighted Awareness Day celebrations, showcasing a noticeable disparity in their frequency compared to the prominence of patient stories and educational material. This discrepancy is likely attributed to the timing of these Awareness Day videos, often concentrated within designated months, weeks, or days dedicated to raising awareness for specific neurocutaneous disorders.

Facebook pages dedicated to neurocutaneous disorders are delineated into four categories: Non-Profit Organizations (NPOs), Support Groups, Private Blogs, and Physician Pages. NPOs and Support Groups were the most prevalent, constituting nearly two-thirds of the page types. A notable observation was the scarcity of physician pages devoted to these diseases. Physician pages have historically functioned as platforms for disseminating educational materials, discussing intricate cases, and extending emotional support among their professional community.^7^ Research studies also showcase the benefits of active participation by healthcare providers on facebook forums in improving patient care and supporting families beyond the hospital walls.^8^ Even with these benefits, it was interesting to see their lack of presence online.^8^ We hypothesize that this may be influenced by concerns regarding patient privacy, the risk of litigation for medical practitioners, and the limited spare time available to physicians. Nonetheless, given the manifold advantages of physician interaction on Facebook, there is a potential opportunity for public policy guidelines specifically tailored for clinicians utilizing social media. Addressing concerns around privacy and litigation risks could facilitate a more inclusive and impactful engagement of physicians on social media platforms, especially within the domain of neurocutaneous disorders.

Top page analysis revealed differences in the types of content being posted. The content on the Neurofibromatosis Network page (NFP) and Tuberous Sclerosis Association (TSA) page most commonly fell under the category of “Page engagement” and “Health education/awareness” which made up 74% and 77%, respectively. The Sturge-Weber Syndrome Awareness day page (SWP)most commonly featured “Personal narratives” from its followers, constituting a substantial 74% of its posts.

Another distinction was observed in the emphasis on research funding and participation. NFP and TSA pages both had 5% of their posts focussing on asking their members to participate in research, whereas SWP pages had 0 mentions of wanting any research participation or funding. These pages’ contrasting natures are influenced by their organizational affiliations. NFP and TSA are non-profit charity organizations, emphasizing fundraising and disease awareness. Conversely, SWP operates as a community-driven platform, with its main focus to cultivate awareness and unity among its members. This focus is evident in the 5% of its content categorized as “other,” concentrating on recruiting research participants and soliciting research funding — a strategy commonly used by researchers studying rare diseases.^2^ These disparities in content and focal points align with established research on disease-specific Facebook pages, underscoring differences in posting strategies and engagement methods across various diseases and even within the same disease.^9^

The most apparent commonality between the SWP, NFP, and TSC included their consistency of posts each week. All three of the organizations had highly active pages with the total number of posts in 2 years being 227, 306, and 366 respectively. People on these pages were always respectful and very encouraging in their comments and engagement on the page. Each page had a slightly different style and themes that they implemented, but they all aimed to create a sense of community and purpose for their members. Another commonality was the public status of these pages, which obviously raised some concerns about patient privacy. Existing research indicates that about two-thirds of such groups are private, resonating with a survey among patients with newly identified or rare genetic findings, where 60% expressed discomfort in sharing information within public groups.^10^ This data suggests a preference for more private settings to discuss specific diseases.^10^ This discrepancy is due to a lack of validation processes for the public accounts which makes it easier for them to amass higher member counts due to ease of access.^10^ Notably, both SWP and NFP occasionally shared social or health advice, often revealing vulnerable information. The TSC group had 0 posts about social or health advice. Patient privacy breaches on social media platforms can carry significant ramifications compared to face-to-face interactions, especially given the absence of privacy protections in public pages.^11^ This underscores the necessity for further research into the advantages and risks associated with divulging personal information on public versus private pages.

## Limitations

This study acknowledges certain limitations, including potential bias in the search results due to Facebook’s algorithm and search functionalities. Additionally, the analysis focused solely on public accounts, potentially excluding valuable information from private or closed groups. Extent of engagement of different types of posts based on likes, comments, and shares remains unknown. The available web pages for neurofibromatosis combined discussions of NF1 and NF2, with a predominant focus on NF1, which limits the comprehensive understanding of the distinct nature and specific needs of NF2. This is a fixed time point evaluation of a single social media platform.

## Conclusion

In conclusion, this study highlights the significant role of social media in supporting patients and families dealing with neurocutaneous syndromes like SWS, NF, and TSC. Personal discussions on these platforms provide a safe space for sharing experiences and insights, and offering valuable support. Our findings underscore the opportunity for greater physician involvement in these online discussions to enhance patient support and disseminate medical advice. Social media can facilitate knowledge sharing and empower patients and families along their medical journey.

## Data Availability

All data produced in the present work are contained in the manuscript

